# Rapid, sensitive and specific SARS coronavirus-2 detection: a multi-center comparison between standard qRT-PCR and CRISPR based DETECTR

**DOI:** 10.1101/2020.07.27.20147249

**Authors:** Eelke Brandsma, Han J.M.P. Verhagen, Thijs J.W. van de Laar, Eric C.J. Claas, Marion Cornelissen, Emile van den Akker

## Abstract

Recent advances in CRISPR-based diagnostics suggest that DETECTR, a combination of isothermal reverse transcriptase loop mediated amplification (RT-LAMP) and subsequent Cas12 bystander nuclease activation by amplicon targeting ribonucleoprotein complexes, could be a faster and cheaper alternative to qRT-PCR without sacrificing sensitivity/specificity. Here we compare qRT-PCR with DETECTR to diagnose COVID-19 on 378 patient samples and report a 95% reproducibility. Patient sample dilution assays suggest a higher analytical sensitivity of DETECTR compared to qRT-PCR, however, this was not confirmed in a large patient cohort. The data showed that both techniques are equally sensitive in detecting SARS-CoV-2 providing an added value of DETECTR to the currently used qRT-PCR platforms. For DETECTR, different gRNAs can be used simultaneously to obviate negative results due to mutations in N-gene. Lateral flow strips, suitable as a point of care test (POCT), showed a 100% correlation to the high-throughput DETECTR assay. Importantly, DETECTR was 100% specific for SARS-CoV-2 and did not detect other human coronaviruses. As there is no need for specialized equipment, DETECTR could be rapidly implemented as a complementary technically independent approach to qRT-PCR thereby increasing the testing capacity of medical microbiological laboratories and relieving the existent PCR-platforms for routine non-SARS-CoV-2 diagnostic testing.

## Introduction

SARS Coronavirus-2 (SARS-CoV-2), the causative agent for coronavirus disease 2019 (COVID-19), emerged in December 2019 in Wuhan, China and caused a pandemic. As of July 19th 2020, over 14 million confirmed SARS-CoV-2 infections and more than 600.000 COVID-19 related deaths have been reported worldwide. To curb this epidemic, effective prevention and control measures including the early identification of SARS-CoV-2 infected individuals, are crucial. Outbreak management is hampered by the high transmissibility and broad spectrum of clinical features of SARS-CoV-2. Severe illness marked by pneumonia, acute respiratory distress syndrome (ARDS) and the need for mechanical ventilation is strongly skewed towards people over 70 years old and those with underlying diseases. Many others experience only mild to moderate symptoms such as fever, fatigue, (dry) cough and/or dyspnoea or do not have complaints at all(1).

Infection surveillance and notification play an important role in outbreak prevention and control. As many infections may go unnoticed, large-scale availability of reliable diagnostic tests also for those with mild symptoms is of critical importance to protect especially those at highest risk of developing severe illness. Accurate monitoring of the SARS-SoV-2 epidemic curve helps estimating future disease burden and serves as an important societal impact parameter for pre-emptive policy making e.g. with regards to the justification of less or more restrictive quarantine measures and prevention of health-care system overflow(2–4). Reverse transcriptase polymerase chain reaction (RT-PCR) is the current diagnostic standard for the detection of SARS-CoV-2. Despite its high sensitivity and specificity, qRT-PCR requires (expensive) specialised equipment, trained staff, and has a relative long turn-around-time (TAT; 2-4 hours). In the Netherlands, the strong dependence on qRT-PCR caused a shortage of reagents and consumables during the pandemic, which limited the test-capacity and resulted in possibly suboptimal outbreak management.

Isothermal reverse transcriptase loop mediated isothermal amplification (RT-LAMP) in combination with Cas12 detection does not need expensive specialised equipment, is highly sensitive and specific, has a short TAT and is easy to implement and therefore could be used as an alternative for qRT-PCR (5,6). This technology is termed DNA Endonuclease-Targeted CRISPR Trans Reporter (DETECTR). The single strand DNA nuclease activity of Cas12 can generate a high-throughput SARS-CoV-2 point-of-care test (POCT) without aspecific amplification as observed with RT-LAMP using intercalating fluorescent dyes or turbidity readouts (5,7), review see (8,9). Since DETECTR depends on both signal amplification by RT-LAMP and reporter degradation after Cas12-dependent amplicon recognition, the assay produces a binary readout and is potentially more sensitive and specific compared to qRT-PCR (5,6). A direct comparison between qRT-PCR and this novel DETECTR assay on a large patient cohort has not yet been performed. In the Netherlands, patients suspected of COVID-19 are admitted under strict isolation procedures to prevent nosocomial transmission of SARS-CoV-2 within the hospital. Unnecessary isolation measures pose a significant burden on the nursing staff as well as on the capacity and costs of the hospital. A rapid highly sensitive SARS-CoV-2 assay, preferably suitable as a POCT, would be of added value for (rapid) clinical decision-making and the optimisation of patient flow within the hospital. In this manuscript we describe the development of an in-house SARS-CoV-2 DETECTR assay, compare its performance with routine diagnostic qRT-PCR on almost 400 patient samples of three Dutch hospitals, thereby providing a first field test of this novel Cas12-mediated SARS-CoV-2 detection tool.

## Materials and methods

All specific information on reagents and relevant concentrations are listed in supplemental tables 1.0-1.6

### RT-Lamp reaction

Primers (supplementary Table 1.1) were dissolved in ultrapure water to a final concentration of 100 µM and prepared in 10x primer master mixes (supplementary Table S1.2). For isothermal amplification, 15 µl of complete RT lamp reaction mix was prepared on ice (supplementary Table S1.3) and incubated with 10 µl of isolated RNA or DNA CTRL plasmid at 62oC.

### RT-(PCR)-Cas12

RNA extracts derived from COVID-19 positive patients were run in a reverse transcriptase (RT) reaction according to table 1.6 and hence amplified with or without PCR. Next, qRT-PCR as well as RT products were incubated with N-gene RNPs and analyzed via HT-detection as described below.

### RNP formation including reporter probe

RNPs were formed by incubating LbCas12 (supplementary Table S1.4) with targeting Guide RNAs in a RNP reaction mix for 30 min at 37oC (supplementary Table S1.5) and subsequently, probe 1,2 or 3 was added in a final concentration of 100 nM (probe 1 and 3) or 500 nM (probe 2).

### High throughput (HT) detection

2,5 µl of RT-LAMP reaction mix was incubated with 22,5 µl of RNP complex containing probe 1 or 3, at 37oC for 10 minutes in chimney multi-well plates covered with seals. Readout was performed after 10 minutes of incubation, unless indicated differently in the figure legends, at 37°C in a Biotek Synergy 2 plate reader using a 485/20 excitation and a 528/20 emission filter.

### lateral chip assay

2 µl of RT-LAMP reaction mix was incubated with 20 µl of RNP complex containing probe 2, at 37oC for 10 minutes. Next, 80 µl NEBuffer2.1 (1x concentrated) was added. Lateral flow strips were incubated for 2 minutes at RT allowing liquid to migrate. Readout was performed visually.

### Double guide approach

Assay was performed as described in HT detection. N-gene Guide 1 and guide 2 were incubated separately as a standard RNP including probe 3. Next, 2 µl of RT-LAMP mix was either incubated with 20 ul RNP mix guide 1, 20 ul RNP mix guide 5, or the combination.

### Dilution experiment

RNA from four DETECR and qRT-PCR confirmed COVID19 patients was diluted in ultra-pure water and 10 µl was used as input for RT-LAMP reaction followed by HT-detection.

### Field experiment: testing of clinical samples

Patient samples from three Dutch hospitals (Hospital A, B and C) were tested for the presence of SARS-CoV-2 RNA with both routine qRT-PCR and DETECTR. The majority of patient samples were nasopharyngeal swabs in transport medium, the remainder were either broncheo-alvealar lavage (BAL) or sputum. Hospital A and C used an in-house one-step qRT-PCR targeting the E-gene of SARS-CoV-2 based on the Cormann primers (10) with the one-step qRT-PCR kit (Roche Diagnostics) or the Taqman Fast Virus 1-step Master Mix (Thermo Fisher), respectively. Hospital B used a commercial dual-target qRT-PCR detecting the S-gene and E-gene of SARS-CoV-2 (Realstar SARS-CoV-2 qRT-PCR, Altona). Hospital C used an integrated system for RNA extraction, amplification and detection of SARS-CoV-2 (GeneLEAD VIII platform, Diagenode Diagnostics), while hospital A and B used two separate platforms (MagNa Pure 96 and Lightcycler 480 II, Roche Diagnostics). All patient samples were spiked with equine arteritis virus (EAV) to detect qRT-PCR inhibition. Not interpretable (NI) qRT-PCR results could result from: delay of the EAV amplification signal with more than 3 cycles, a weak non-exponential EAV or SARS-CoV-2 amplification signal, or EAV or SARS-CoV-2 amplification signal with a strongly reduced maximum fluorescence. All patient samples were anonymized according to EU-privacy regulations and were used in accordance with the declaration of Helsinki, approval to use material was obtained via internal hospital ethic boards.

### Statistics

All data was first tested for normality by the Shapiro Wilk test (p=0,05). Data with a gaussian distribution was analyzed with an unpaired two-sided student’s t-test in case of the comparison of 2 samples or an one-way ANOVA with a Dunnett’s post-test in case of 3 samples or more. All statistics were analyzed in Graphpad Prism version 8.0.2.

## Results and discussion

Both (RT-)LAMP and Cas12-RNPs can be used to detect RNA/DNA, while the combination potentially increases sensitivity and specificity (6). We compared the sensitivity of RT-LAMP, RT-Cas12-RNP and DETECTR (combination RT-LAMP/RT-Cas12-RNP, Figure 1A; supplemental figure 1A-C). We show that using solely RT-LAMP (figure 1B) or RT-Cas12-RNP (figure 1C and Supplemental figure 1D) did not match the sensitivity of DETECTR (figure 1D, 1E). Of note, the limit of detection (LOD) for RT-LAMP was similar to previously reported (11). RT followed by Cas12-RNP was not sufficient to detect SARS-CoV-2 RNA in samples with high SARS-CoV-2 viral load (qRT-PCR, Cq-value<20). This emphasizes the importance of a separate amplification step (PCR or LAMP) prior to Cas12 detection (Figure 1C and Supplemental figure 1D). The added value of Cas12-RNP shows in the improved signal-to-noise ratio, which eases interpretation, compared to RT-LAMP alone (figure 1E). Interestingly, Cas12-RNP by itself also displays a dependency on target concentration (figure 1C; supplemental figure 2A). This suggests that the RT-LAMP reaction is required to allow sufficient amplification of Cas12-RNP target DNA to allow efficient probe degradation. To investigate the effect of probe length on assay performance, we tested a wide range of SARS-CoV-2 N gene DNA (range 10-7 to 10-16M) using probes of 8 and 12 nucleotides (nt). The use of a 12 nt probe increased the signal to noise ratio but not the sensitivity of the test (Supplemental Figure 2A-B). The plateau of the fluorescent signal using DETECTR is reached after 10 minutes. However, >75% of the maximum fluorescence is reached within 5 minutes, suggesting that the assay can be performed faster if required (Supplemental Figure 2C). Longer incubation does not increase the fluorescent signal (Supplemental Figure 2D). However, plates can be re-measured or stored for at least three days without significant loss of signal when stored at room temperature in ambient light. In conclusion, our DETECTR data confirm short turn-around-times (<30 minutes including RT-LAMP), signal robustness and ease of result interpretation.

**Figure 1:**
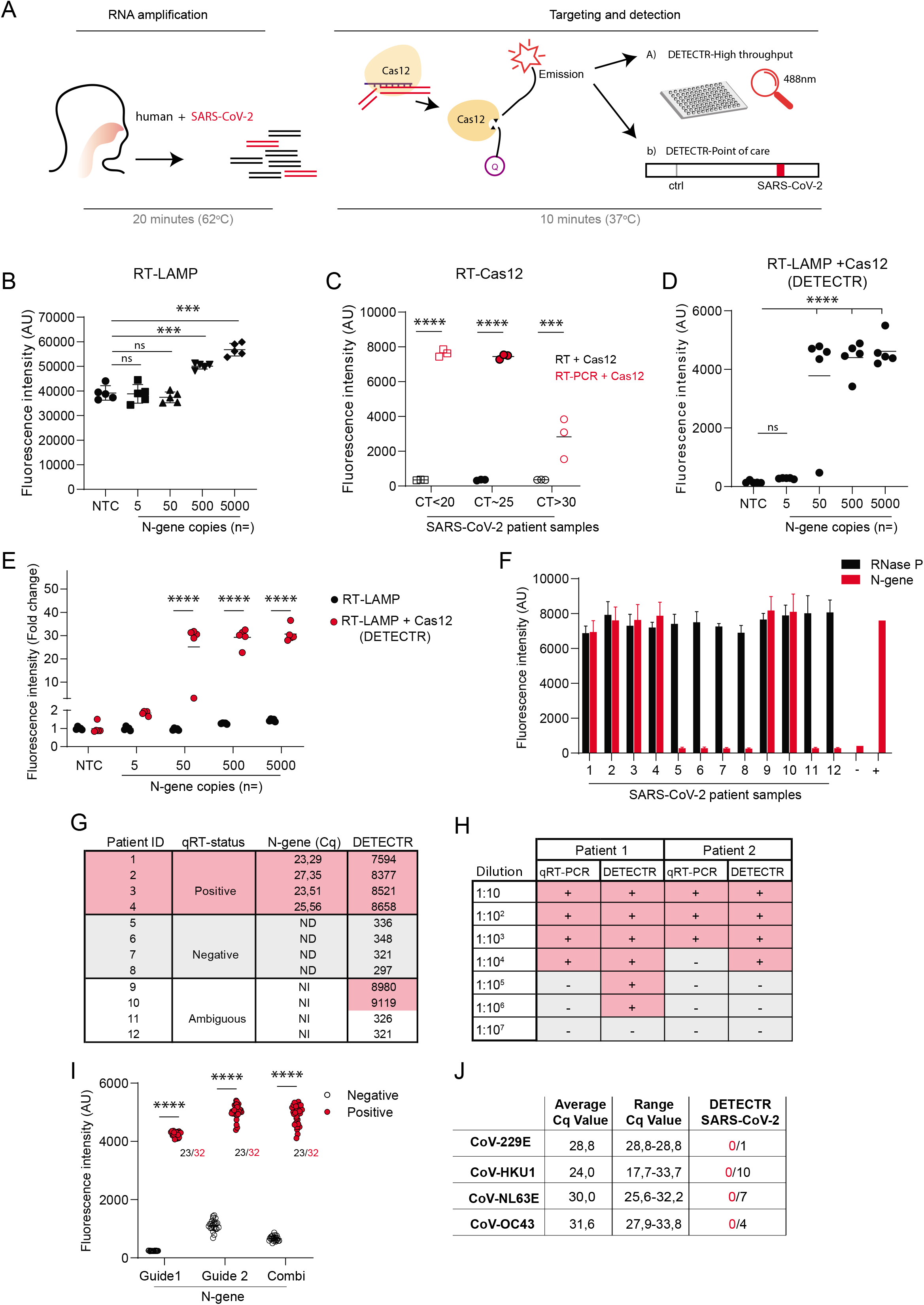
RT-LAMP/Cas12 is a specific and sensitive test to detect SARS-CoV-2. A) Graphic representation of DETECTR assay: RNA is converted to cDNA and amplified in one reaction mix for 20 minutes using RT-LAMP. Next, Cas12 RNPs are added that recognize and cleave SARS-CoV-2 amplified products leading to activation of Cas12. Activated Cas12 destroys the single stranded linker DNA between quencher and probe leading to fluorescence that can be detected on indicated platforms. B-E) Specific N-gene plasmid copy concentrations (B,D) or confirmed RNA from SARS-CoV-2 positive individuals with indicated range of qRT-PCR cq-values (C) were run in a RT-LAMP reaction (B), RT-Cas12 (C) or RT-LAMP-Cas12 (DETECTR, D) assays using an amplicon within the N-gene and a gRNA annealing to that amplicon (supplemental figure 1). Note that whereas the RT-LAMP reaction results in a fluorescence signal proportional to the input (B), the combination of RT-LAMP/Cas12-RNP results in a binary test outcome with a high signal to noise ratio (24 times on average) due to the degradation of the reporter probe, which depends on the induced nuclease activity of Cas12 (D). E) shows the background to signal ratio fluorescence fold change normalized to the negative control. F) a cohort of 12 patient RNA isolates, including 4 RT-PCR positive, 4 RT-PCR and clinically negative and 4 NI were screened with DETECTR (the convention of a NI (not interpretable) result can be found in material and methods). Bars represent the average of a duplicate and error bars the SD (N-gene (red) and internal control RNAseP (black)). G) shows the comparison of qRT-PCR result and DETECTR fluorescence signal (red positive samples; grey negative samples; white NI. H) Log scale dilution of positive samples from figure F were again tested with qRT-PCR and DETECTR in indicated log dilutions were + indicates a positive result and – a negative test result. (I) Two gRNAs with distinct annealing sites were tested in the DETECTR assay. The dot plot shows the fluorescence signal of positive (red) and negative samples (black). Numbers indicate the number of negatives and positives samples analyzed. J) A collection of non-SARS-CoV-2 corona strains samples confirmed by qRT-PCR were found to be negative using SARS-CoV-2 specific DETECR. ***=p<0.001; ****=p<0.0001 using ANOVA and Dunnett’s post-test (B-E) or two-sided unpaired T-test.

In a pilot experiment we blindly tested a small cohort of patient samples including four positive, four negative and four samples with not interpretable (NI) qRT-PCR results. SARS-CoV-2 RNA was detected in all 4 qRT-PCR positive samples plus 2 qRT-PCR NI samples (Figure 1F, G). Human RNAse P RNA, used as an internal control, was detected in all 12 samples. Hence, DETECTR results were consistent with qRT-PCR, and provided a clear-cut positive (n=2) or negative (n=2) test result for the samples with NI qRT-PCR results. The analytical sensitivity of DETECTR was compared to qRT-PCR using log-scale dilutions of SARS-CoV-2 RNA extracted from patient samples. DETECTR proved 10-100 times more sensitive in 3 out of 4 experiments (Figure 1H, supplemental figure 2E and F). Of note, the observed analytical sensitivity of both tests does not necessarily equal their clinical sensitivity as (potential) inhibitory factors present in patient material have also been diluted. As the Cas12-RNP complex is single nucleotide sensitive (12,13), mutations within the gRNA recognition site may prevent Cas12 detection. Using a dual target approach with gRNAs that anneal to distinct parts of the RT-LAMP generated amplicon could prevent escape from Cas12 detection (supplemental figure 1B). DETECTR results with gRNA1, gRNA2 and combined gRNA1/gRNA2 yielded similar results (Figure 1I). As the risk of aberrant viral variants increases with the ongoing worldwide epidemic, the use of multiple gRNAs is highly recommended. Strong homology within the N-gene of SARS-CoV-2 and other human coronaviruses may compromise the specificity of DETECTR. N-gene homology with other human coronaviruses varies between 50.1% and 88.2%. The highest concordance is seen with SARS-CoV-1, with a maximum homology of 86.7% in the regions used for the development of RT-LAMP primers and Cas12 gRNA recognition sites. We analyzed 22 samples of patients infected with other human coronaviruses; 22/22 samples tested negative for SARS-CoV-2 and positive for the RNase P housekeeping gene with DETECTR suggesting a specificity of 100% (Figure 1J).

Finally, we tested our DETECTR assay on 378 patient samples derived from three hospitals in the Netherlands. The cohort consisted of RNA extracted from clinical samples of patients that were diagnosed SARS-CoV-2 positive or SARS-CoV-2negative based on routine qRT-PCR. Our DETECTR assay showed 94.9% (+/- 1.8%/0.8%) concordance with qRT-PCR (figure 2A, 2B), with minor differences between the three centers (Supplemental figure 3; A=94.1%; B=96.7%; C=94.7%). DETECTR positive but qRT-PCR negative samples (n=10) were mainly found in center A (n=9); all 9 samples from this hospital showed a SARS-CoV-2 band pattern on gel suggesting they were missed by qRT-PCR (supplemental figure 3D). The DETECTR+/PCR-negative sample of center B was later confirmed SARS-CoV-2 positive by Center B, albeit with Cq-value >35. In total, we found 11 PCR+/DETECTR-samples, 7/11 samples had Cq-values >30 (figure 2B; supplemental Figure 3A-C), but the other four had Cq values of 20,74; 29,78; 29,28 and 28. Re-analysis with an alternative gRNA that anneals to a different part of the N-gene (supplementary Table S1.1) did not yield positive test results, making a single nucleotide gRNA escape mutation less likely. Alternatively, the clinical sensitivity of DETECTR could be lower compared to qRT-PCR. Despite its higher analytical sensitivity (Figure 1F, 1H), the matrix of clinical samples could have a more profound inhibitory effect on DETECTR technology. However, patient samples were selected based on qRT-PCR results with an overrepresentation of samples with high Cq-values (>30) which could indicate a higher clinical sensitivity for qRT-PCR. Almost an equal amount of patient samples were SARS-CoV-2 positive with DETECTR and SARS-CoV-2 negative with qRT-PCR, of which the viral load can be expected to be around the LOD of qRT-PCR. However, DETECTR confirmed the presence of SARS-CoV-2-RNA in 9/19 patient samples with a not interpretable qRT-PCR result (figure 2C), indicating a higher specificity of DETECTR. Altogether, the overall concordance of around 95% in clinical sensitivity, shows that DETECTR can be used as a specific, fast and reliable technique for patient samples.

**Figure 2.**
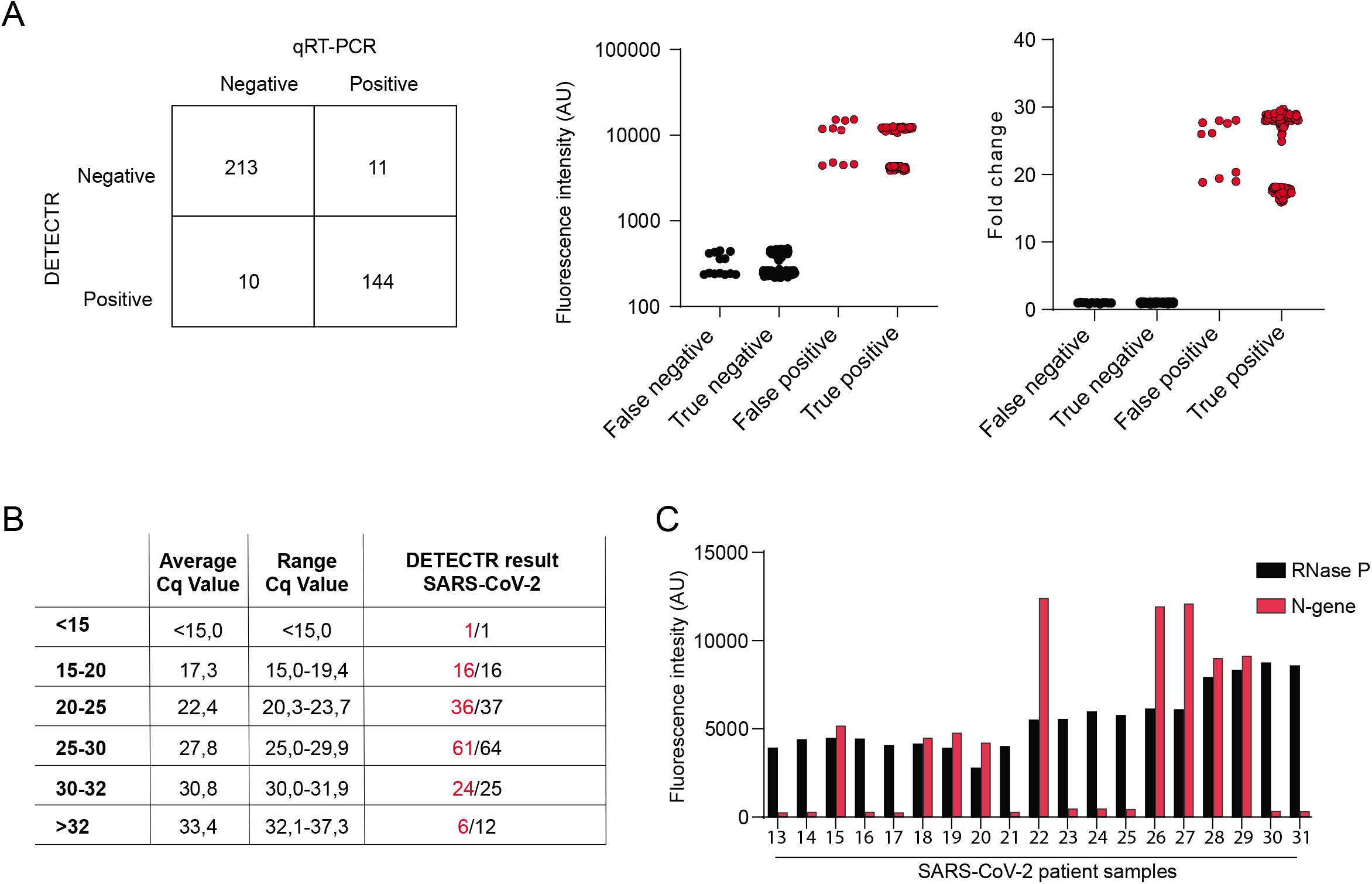
High level concordance between qRT-PCR and DETECTR. A) 378 qRT-PCR confirmed SARS-CoV-2 tested samples from different centers were compared to the results obtained using DETECTR. The matrix displays the results from the DETECTR assay (vertical) compared to the qRT-PCR results (horizontal). The graph shows the fluorescence intensity (left graph) and fold change fluorescence signal normalized to a negative control (right graph) with each dot representing a DETECTR test on RNA from a different individual. B) Subclassification based on qRT-PCR cq-values compared to DETECR result. C) DETECTR on 19 samples that gave an NI results by qRT-PCR (convention of NI can be found in Material and Methods). Orange bars: N-gene DETECTR; black bars: RNaseP control DETECTR.

Most DETECTR results were obtained using a high throughput 96/384 wells spectrophotometer to detect the cleaved fluorescent probe. A major advantage of DETECTR is that it can be used as an individual POCT using lateral flow strips for read out. Individual lateral flow results (n=40) were 100% concordant with the high throughput results (supplemental figure 4). To confirm robust signals in ‘difficult’ clinical samples, we analysed 8 samples with not interpretable qRT-PCR results using spectrophotometric and lateral flow detection. Again, fully concordant results: SARS-Cov-2 positive (n=4) and SARS-CoV-2 negative (n=4) (Supplemental Figure 4C). The binary readout is easy to interpret, irrespective of readout method or Cq-value. Therefore, DETECTR POC tests could be used in low-resource countries/regions or as a fast and reliable equipment independent confirmation test to confirm ambiguous qRT-PCR samples.

In summary, here we compared DETECTR with qRT-PCR for SARS-CoV-2 diagnosis in a large patient cohort over multiple hospitals and report a 95% accordance. These data are in line with a recently published study where only a small cohort (83 samples) was tested derived from a single hospital. In addition, our data suggest that a 12nt probe is superior over a 8nt probe and we suggest to use a double guide approach to prevent escape from DETECTR due to variations within amplicons. Overall, DETECTR has comparable sensitivity and superior specificity to qRT-PCR. Our results show that DETECTR represents a reliable, cheap, fast and technically independent alternative to complement qRT-PCR platforms. The low-demand on facility equipment, especially concerning the POCT, makes DETECTR especially suitable for resource low countries/regions. However it must be noted that presently DETECTR is a three step reactions involving separate RNA isolation, RT-LAMP amplicon amplification and Cas12 mediated reporter degradation. The latter has to be considered a step back in comparison to qPCR, where post amplification handling, a major risk in causing false positive results by contamination, could be removed from the workflow. Further research should focus on integrating all DETECTR steps, including RNA-isolation, into the same reaction tube without post amplification processing. In the current study, the extracted RNA used as input for qRT-PCR was also used for DETECTR. Of note, onestep RT-LAMP approaches including various RNA extractions have been developed, e.g. for Zika virus(14–16), and compatibility with DETECTR will need to be determined. Importantly, as detection is not compromised upon diluting patient material 10-100 times, the technique may allow the implementation of pooled sample approaches in low-prevalence regions/countries significantly increasing testing capacity (e.g. 20 samples without loss of detection). However, it must be noted that in this patient cohort DETECTR and qRT-PCR were performing on parity. Importantly, once implemented the suggested approach can be easily diverted to screen other existing or emerging pathogens or any other platform that requires identification based on specific DNA/RNA (6,12,13). The DETECTR test helps to optimize diagnostic strategies for both bedside and high-throughput settings leading to an increase in testing capacity and improved diagnostic evaluation, ultimately leading to better determination of endemic progression facilitating governmental policy decisions.

## Data Availability

All used data is reported in the paper. No complex datasets have been used within this paper.

## Acknowledgements

This study was supported by a NWO-ZONMW “creative solutions to battle COVID-19” grant from the Dutch government (No.5000.9954). We would like to thank the individuals part of the studied cohort.

## Authorship contributions

EB and HV performed the RT-LAMP, DETECTR assays. MC, TvdL, EC and JS collected the cohort material, isolated RNA and performed the qRT-PCR on validated platforms. EvdA supervised the study. All authors contributed in writing the manuscript, which was critically reviewed by all authors.

## Figure legends

**Supplemental figure 1.**
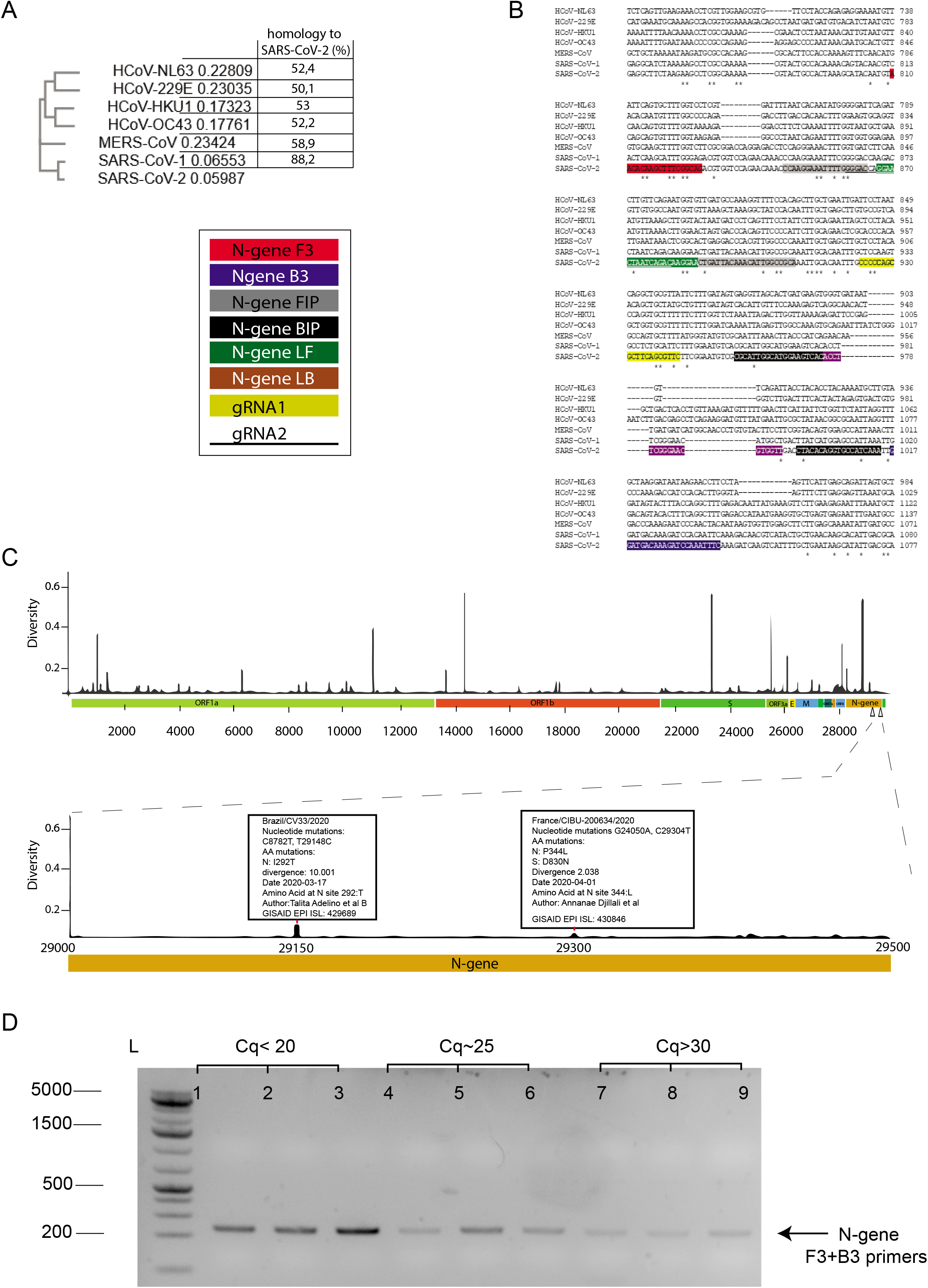
A) The N-gene sequence of specific Corona virus variants as indicated was aligned using clustalOmega (17) and the evolutionary phylogenetic tree (A) and alignment (B) is shown. B) Indicates the N-gene specific LAMP primers and gRNAs used within the study. The colors are used to indicate the location within the N-gene sequence in. C) Upper part shows the complete SARS-CoV-2 genome and lower zoomin the amplicon location within the N-gene amplicon (29071-29278; MT628280.1). The y-axis indicates the global variant frequency distribution (over 3000 different SARS-CoV-2 genomes included; GISAID database (18), of specific SARS-CoV-2 variants (black bars) of which two mutations are highlighted. The variations found in the GISAID repository within the gRNA targeted sequence are indicated in bold within (B). (D) Agarose gel that belongs to Figure 1B showing the RT-PCR products that correlate to the expected size of 220 bp.

**Supplemental figure 2.**
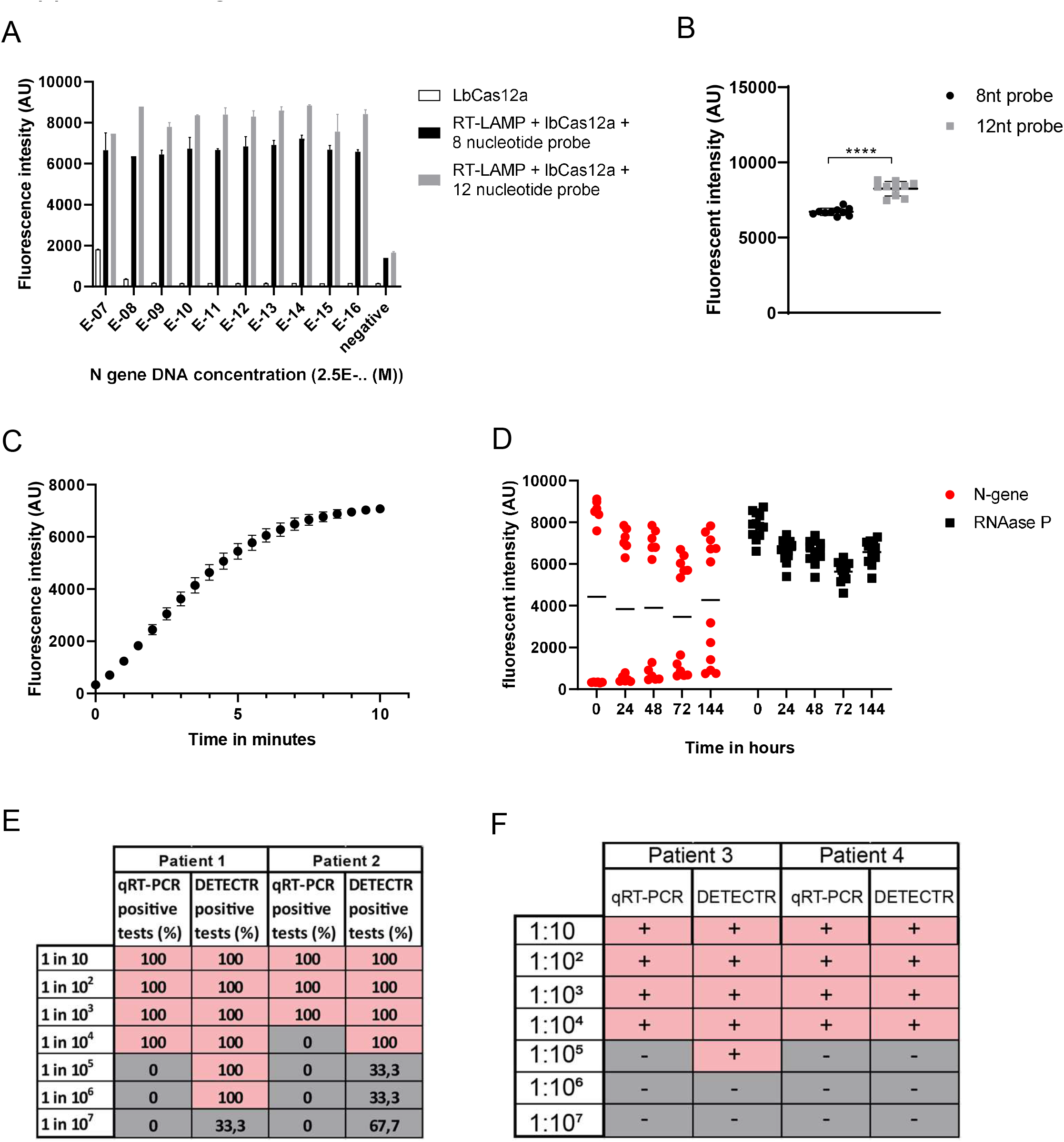
A and B) Various concentrations (x-axis) of N gene plasmids were tested in a LAMP-Cas12 or Cas12 only assay in presence of an 8nt or 12nt length probe (supplemental table 1.1). Bars represent the median readout of a duplicate and the error bars show the standard deviation (A). B) Fluorescence intensity for the 8nt (black) and 12nt (grey) probe is shown for the 8 samples in A (paired two tailed student t-test, ****=P <0.0001. C) isolated RNA from SARS-CoV-2 positive samples was subjected to the DETECTR and the fluorescent intensity over time was measured as a readout for probe degradation. The dots represent the mean and error bars the standard deviation (RNA from two individuals in triplo). D) From the cohort, 6 positive and 6 negative samples were subjected to DETECTR and analyzed after 10 minutes (t=0) and re-measured on the indicated time points (hours). Note the stability and robustness of the signal to noise ratio. E) Log scale dilution of positive samples belonging to figure 1H of positive individuals compared to qRT-PCR. The percentage indicates the detection in a triplicate experiment for both qRT-PCR and DETECTR. Note that DETECTR is still performing on a “hit and miss” basis in the highest dilutions. F) A comparison between qRT-PCR and DETECTR on two additional positive samples. Importantly, the DETECTR in (F) was performed on frozen RNA while the qRT-PCR in (F) was performed on freshly isolated RNA. Freeze thaw samples lead to loss of RNA, interfering with detectability.

**Supplemental figure 3.**
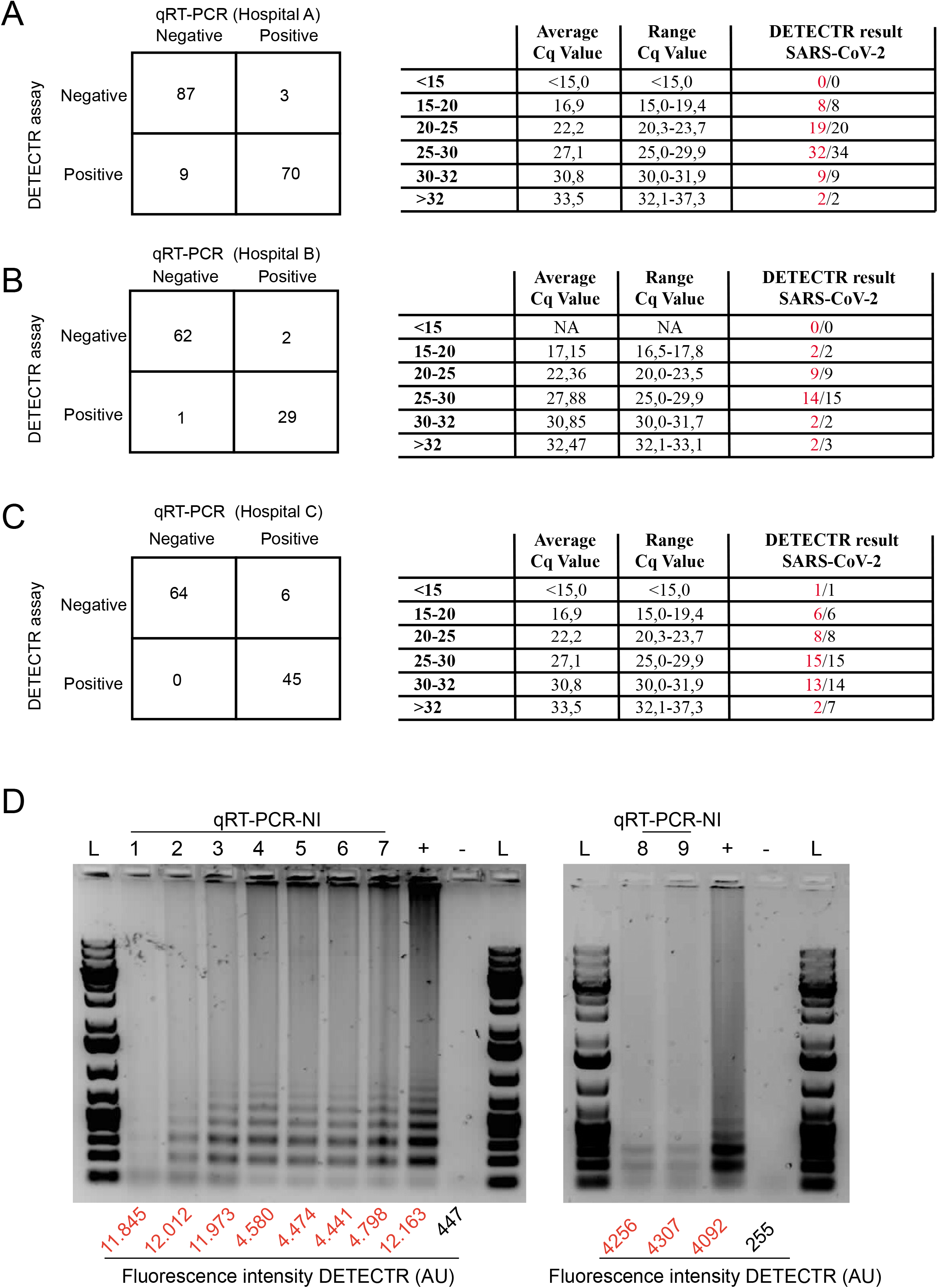
The comparison between DETECTR and RT-PCR as a function of qRT-PCR cq value shown for the individual centers (A, B and C). The matrices depict the DETECTR-assay (y-axis) vs the qRT-PCR (x-axis) from center A, B or C as indicated. The tables show the results ranked by increasing qRT-PCR cq value for the individual centers as indicated. D) RT-LAMP with SARS-CoV-2 N-gene specific primers was conducted on all 9 false positive patients (FP) from center A that were positive by DETECTR. The agarose gel shows the characteristic clear step-wise incrementing RT-LAMP loop product bands for all 9 false positive as well as for a positive control (lane 9, left gel; lane 4, right gel)). Note that these bands were absent upon performing the RT-LAMP on RNA from a confirmed negative sample (lane 10, left gel; lane 5; right gel). RT-LAMP results left gel: 1; gene ruler 1 kb plus Basepair ladder (Bp), 2; FP 1, 3; FP 2, 4; FP 3, 5; FP 4, 6; FP 5, 7, FP 6, 8; FP 7, 9; positive patient sample, 10; negative patient sample, 11;Bp; right gel: generuler 1 kb plus Basepair ladder (Bp), 2; FP 8, 3; FP 9, 4; positive patient sample, 5; negative patient sample, 6;Bp. Numbers at the bottom of the gel indicate the DETECTR fluorescence intensity.

**Supplemental figure 4.**
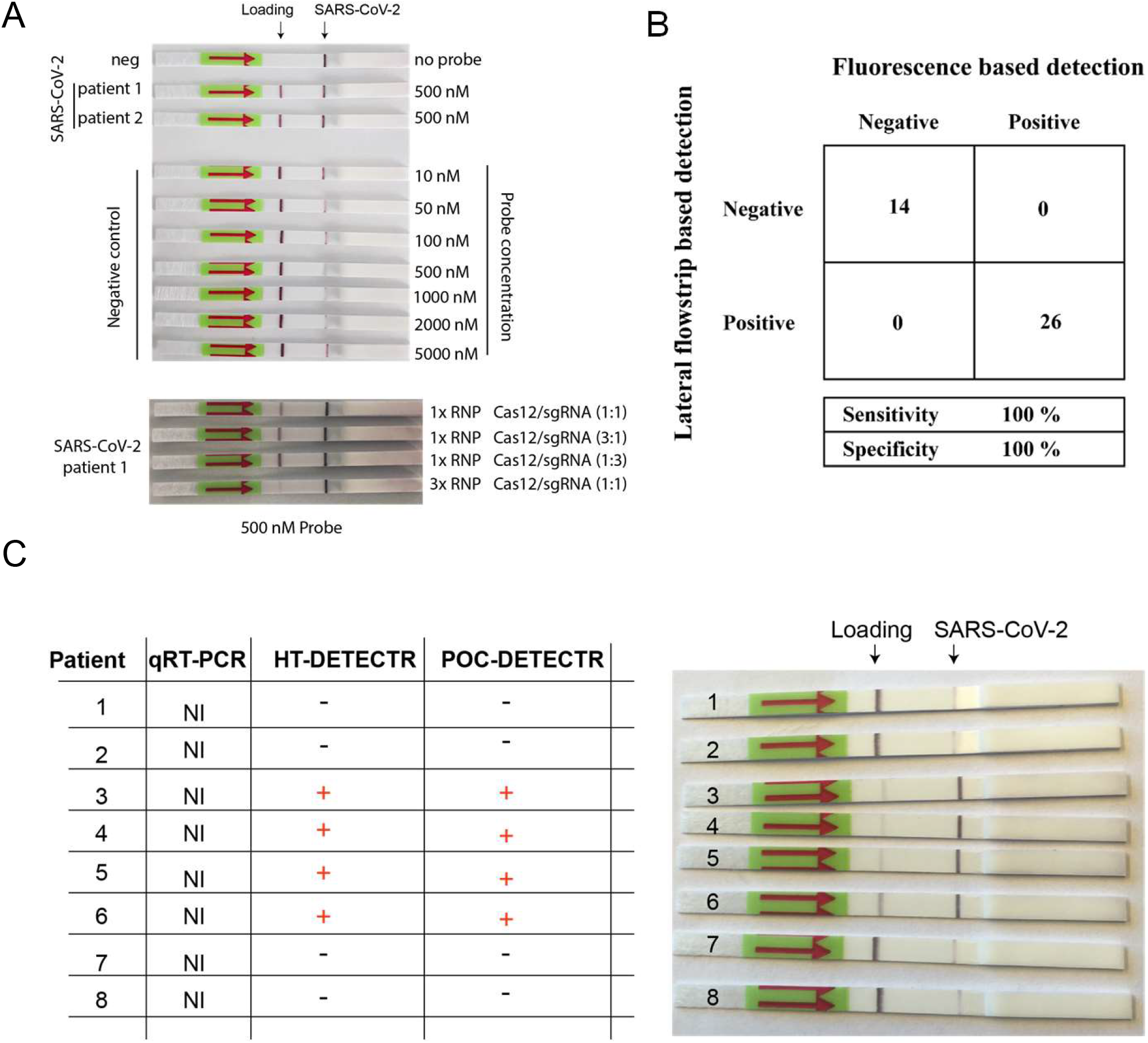
A) shows representative pictures of the lateral flow assays performed after RT-LAMP-Cas12 using probe 2 (table S1.1). Distinct probe and RNP concentrations were tested. 500 nM is the most optimal probe concentration. No differences were observed between different Cas12/sgRNA ratio’s, however, 3x higher dose of RNP leads to substantial more cleaved SARS-CoV-2 cDNA (lower strips). Note that probe degradation in positive samples leads to the appearance of a second dark black band (indicated as SARS-CoV-2). The first left black band serves as a probe loading control (loading). B) matrix shows complete correspondence between the fluorescence plate reader results using probe 1 (table S1.1; x-axis) and the lateral flowstrip results (y-axis; 40 samples). C) 8 samples with a qRT-PCR NI status were analyzed with HT-DETECTR and POCT-DETECR, both DETECTR tests are in agreement.

